# Mapping Of The Sino-European Science And Technology Collaborations On Personalized Medicine: An Updated Overview

**DOI:** 10.1101/2024.06.27.24309578

**Authors:** Letizia Pontoriero, Andrea Mazzoni, Giovanni de Santis, Matteo Gentili, Ejner Moltzen, Sabine Puch, Carolin Lange, Andrea Frosini, Gianni D’Errico

**Affiliations:** Fondazione Toscana Life Sciences, Firenze, 53100, Italy; Erre Quadro s.r.l., Pisa, 56126, Italy; Innovation Fund Denmark, Aarhus, 8000, Denmark; German Aerospace Center, Project Management Agency, European & International Cooperation, Asia, Oceania, Bonn, 53227, Germany

**Keywords:** Sino-EU PerMed, ICPerMed, EPPerMed, Patent Mapping, Scientific Mapping, Personalised/Personalized Medicine, Deep Phenotyping, Omics Sciences, Big Data, Machine Learning Techniques

## Abstract

**Aim:** Personalized medicine is part of the future frontier of public health and precision healthcare systems have been implemented for years, both within Europe and beyond. To establish the state of the art of Sino-EU science and innovation in Personalized medicine, we have mined the major dedicated databases globally. Here we present the updated mapping on the Sino-EU collaborations. Patents, scientific publications and preprints related to Personalized medicine have been mapped and analyzed after being extracted through databases mining. The integration of the previous mapping provides a more complete overview, which does not show relevant variations, confirming previous trends. In this work we complete the mapping by providing a digital tool for consulting the various data collected.

## Introduction

While a universally accepted definition is lacking, Personalized Medicine (PM) is most described as a “*a medical model using characterization of individuals’ phenotypes and genotypes (e.g. molecular profiling, medical imaging, lifestyle data) for tailoring the right therapeutic strategy for the right person at the right time, and/or to determine the predisposition to disease and/or to deliver timely and targeted prevention*”.

The definition was offered by the Horizon 2020 Advisory Group and adopted by the European Union (EU) Health Ministers in their *Council conclusions on personalized medicine for patients*, published in December 2015[1] .

The very concept of PM took hold decades ago[2] underpinned by the expectation that emerging technical capabilities, such as predicting health risks, monitoring disease progression and predicting responses to treatments, could unveil personalized and preventive approaches to healthcare. Thus, the idea of PM has undergone a gradual evolution expanding to become synonymous with personalized healthcare[3]. The urgency to transform the current healthcare paradigm from a reactive, disease-focused model to an individualized, anticipatory, and preventive method, that puts patient’s needs first, is indeed evident nowadays.

Health systems globally aim to adopt an increasingly personalized approaches to medicine, and, in this context, the EU has been and is actively working towards this realization, encouraging collaborations and projects on this topic. The International Consortium for Personalized Medicine (ICPerMed)[4] and the recently launched largest PM initiative, the European Partnership for Personalized Medicine (EP PerMed) [5], represent two significant initiatives which support the implementation and transnational development of personalized medicine approaches, and its focus is not limited to European borders. It is pivotal to assess the approach to PM in other countries, and to adapt to the regional and national conditions, where different policies and regulations apply, in order to succeed globally in implementing the PM-based approaches in healthcare. One size does not fit all to.

Through cooperation, knowledge sharing and collaboration, the global health architecture can accommodate the strategies required to enable the development and implementation of PM-based approaches for the benefit of patients and citizens, taking into account cultural and societal differences [6]. It is thus fundamental to explore the extra-EU healthcare landscape, and to identify and encourage possibles areas of collaboration.

China is one of the most important global players in life science research and innovation, which in recent years has consolidated its leadership in health technology and has long been working on large-scale PM initiatives thanks to its formidable genome sequencing capability and strong government support [7].

With the Belt and Road Initiative (BRI) [8],in particular, China has declared its willingness to actively participate in finding the best solution to the main global challenges. Health cooperation, in particular, was boosted and gained significant attention. For instance, China signed a memorandum of understanding with WHO on health cooperation in BRI partner countries and also signed health cooperation agreements with over 160 countries and international organizations.

The EU and China share a significant and enduring relationship, being among the world’s largest economies and traders. Although both sides are committed to a comprehensive strategic partnership, there is a growing realization in Europe that the challenges and opportunities presented by China have evolved [9].

The actual complex global scenarios hampered the possibility of interaction between the two regions. Nevertheless, this does not dampen the desire for collaboration, that must be supported.

The post-COVID-19 era has shown the need to deepen and improve China-Europe health cooperation, emphasizing various aspects such as improving health policies, strengthening cooperation in disease prevention and control, and promoting training and capacity building. In particular, the potential for deeper cooperation between China and the EU in PM is very high.

In this framework, the project ’Widening Sino-EU policy and research cooperation in Personalized Medicine’ (Sino-EU-PerMed)[10] aims to align with ICPerMed by linking strategies and activities with Chinese stakeholders. Sino-EU-PerMed started in 2020 and coordinated different activities fostering better mutual understanding of PM research and innovation as well as cultural aspects including related ethical and legal issues.

As part of the project, a thorough analysis of existing literature, encompassing scientific papers and patents on PM was conducted. The aim was to provide a comprehensive, transparent, and objective overview of the R&I landscape within PM in China and Europe and establish the current level of collaboration in science and technology through comparison. This encompassed identifying key research areas, innovation initiatives, collaborations with European researchers, and elucidating the support mechanisms for PM through research programs.

The work was presented in the research article by Romagnuolo et al. “Sino-European science and technology collaboration on personalized medicine: overview, trends and future perspectives”[11].

The Sino-EU PerMed project has updated the 2010-2020 mapping. This now covers information on technological and scientific collaborations in PM and its development from 2020 to December 2022, including preprints as a new category. As additional element of innovation, in consideration of the importance of preprints repositories during the pandemic, we have included these databases in the mapping updates.

The mapping has been recently updated using the same workflow, and the consolidated data is shared through an interactive web platform accessible via the project website.

## Mapping methodology: Patents

### Tailored vocabulary and terminology for databases mining

PM remains a comprehensive term [12] and this led to define specific vocabulary.

Consequently, the mapping process requires the identification of suitable keywords and precise definitions formulating queries when mining the selected databases.

The **definitions** are:

1. ***Personalized medicine:*** a medical model using characterization of individuals’ phenotypes and genotypes (e.g. molecular profiling, medical imaging, lifestyle data) for tailoring the right therapeutic strategy for the right person at the right time, and/or to determine the predisposition to disease and/or to deliver timely and targeted prevention (European Council Conclusion 2015/C 421/03).
2. ***Precision medicine:*** treatments targeted to the needs of individual patients based on genetic, biomarker, phenotypic or psychosocial characteristics that distinguish a given patient from other patients with similar clinical presentations” [13] Focus on process and used data: precision medicine as “a model that integrates clinical and other data to stratify patients into novel subgroups.
3. ***Personalized health care & Precision public health:*** the application of clinical know-how, concepts of systems medicine, and PM technologies to improve health and minimize disease.

The **keywords** to be used within the searches are *personalized medicine, personalized medicine, precision medicine, preventive medicine, predictive medicine, systems biology, systems medicine, stratified medicine, targeted therapy, tailored treatment/therapy, deep phenotyping, omics sciences, big data, machine learning techniques, theragnostic, oncogenomic, pharmacogenomics, patient-specific treatment, individualized therapy*.

The **established goal** of PM can be:

- Diagnostics
- Design of patient-specific therapy/treatment strategy
- Forecast of success of therapy/treatment strategy

In addition, it was assumed that the **tools of PM** are genetic analysis, statistical analysis, patient-specific treatment, patient-specific drug.

To exclude inconsistent results that deviate from the definitions of PM, the research process was meticulously refined based on assumptions regarding the characteristics of PM and clearly delineating what falls within its scope and what does not.

**PM is:**

- Drugs, devices, methods for patient-tailored treatment/therapy
- Procedures using an individualized approach based on genetic evaluation and treatment
- Treatment strategies based on individual data of patient genotype, phenotype, lifestyle
- Methods for synthetizing or designing customized and patient-specific drugs
- Methods for predictive medicine (e.g. statistical method for predicting life expectation, therapy response based on patient genotype, phenotype, patient lifestyle stratification)

**PM is not:**

- Customized prosthesis/implants
- Medical instrument designed for specific group of patients (e.g. obese patients)
- Diagnostic, therapeutic and prophylactic approaches for a specific disease (and not for a specific patient)
- Genetic analysis for general purposes

Other fields excluded are:

- Veterinary
- Personalized nutrition plans

### Patents database mining

To identify PM-related patent documents, it was adopted the functional approach developed by Bonaccorsi et al.[14]. While being pivotal for the patent analysis and their widespread applications, the current patent classification system has limitations. Specifically, it struggles to fully capture the interdisciplinary nature of inventions. Bonaccorsi’s approach proposes integrating existing classification schemes with a comprehensive functional classification based on the primary functions performed by a technology. This approach aims to overcome the limitations of traditional classification systems by providing a more generalized and abstract representation of functions. This was of particular importance when classifying patents in categories not published under Cooperative Patent Classification (CPC) or International Patent Classification (IPC) [15]. Indeed, relevant patents are occasionally published under different classifications from the initial patent application. Traditional queries often fail to detect them due to overreliance on CPC/IPC patent classification or the use of domain-dependent keywords.

Furthermore, patents may articulate a concept through periphrastic constructions rather than using terms directly associated with PM, and traditional semantic approaches may pose a challenge in this complex, multidisciplinary field. To solve this issue, the functional approach mentioned above was applied, with the aim of iteratively improving and refining the definition while improving the recall of the patent set.

The operational workflow created for the initial mapping was retained and depicted in Figure 1. The latter provides a visual representation of the step-by-step approach employed for patent mapping, offering a comprehensive overview of the systematic process followed in mapping activities.

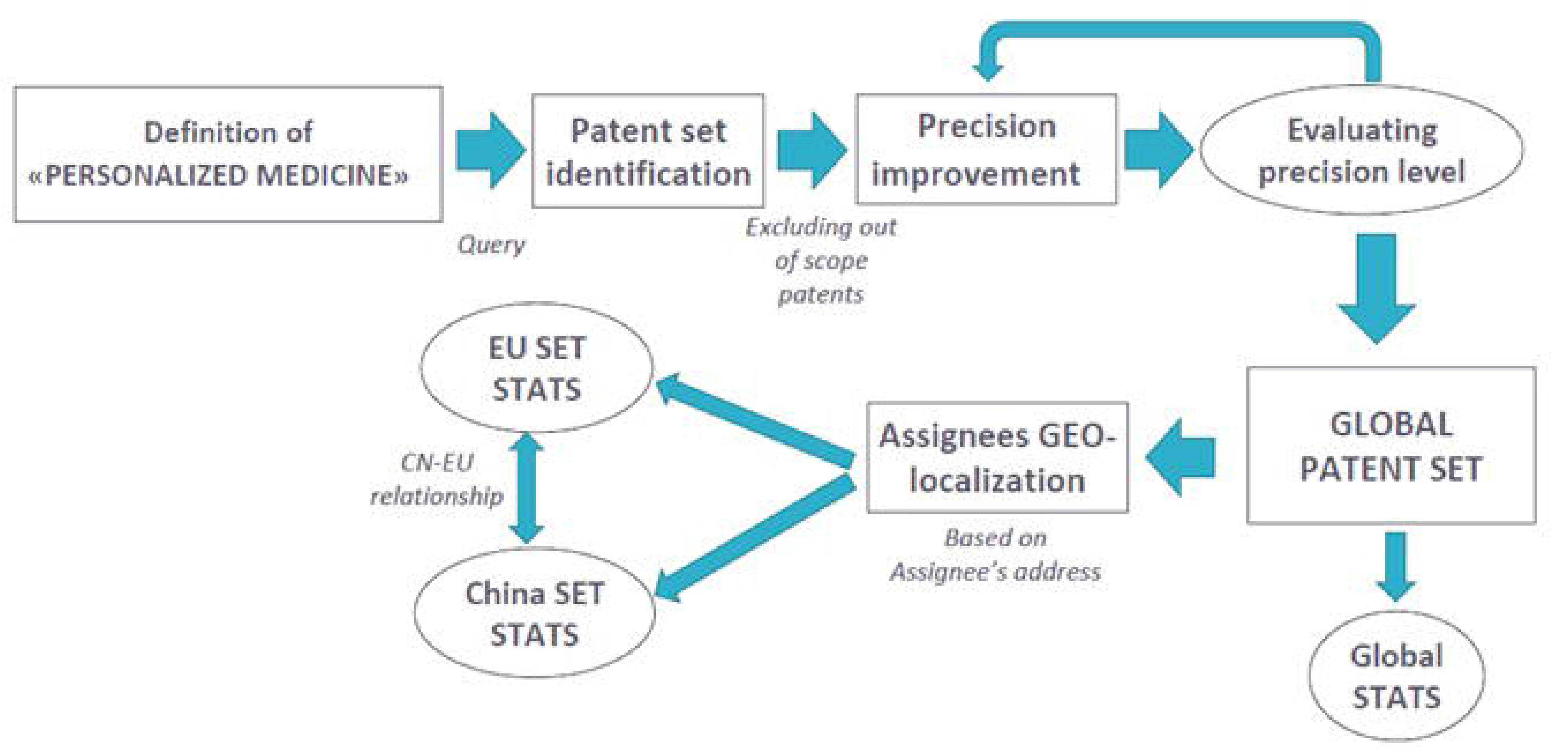

The information provided on national patent validations within the dataset allowed the dataset to be clustered into two distinct parts: one for Europe and one for China. Specific data variations between these locations were presented along with the overall information of the dataset.

### Global set analysis

The presented analysis has been carried out using documents from the Erre Quadro proprietary database, based on the data provided by the PATSTAT service based on European Patent Office (EPO) database[16].

In some cases, data supplied by Chinese Patent&Trademark Office to EPO are partially omitted. For those documents, the geographic location of the address is retrieved using other proprietary tools developed by Erre Quadro. However, for a restricted number of documents, it is not possible to obtain a reliable geographical location. For such reasons, the information based on geographical localization retrieved in this analysis can be underestimated.

The patent analysis is always affected by the blind period: usually a patent application is published several months after the filing and could be unavailable in the patent databases. Updating the analysis after 2-3 years can overcome this issue. With respect to the previous analysis, the presented outcomes overcome part of the blind period from the previous mapping, giving information on patents filed by 2021. However, they are limited to 18 months before 31 December 2022.

Table 1 compares updated data with the previous patents’ mapping outcome. The parameters are:

- **Patent families**, namely sets of patents or patents application filed and extended in various countries which cover the same or similar inventions and are disclosed by common inventors[17]. Having the same priority date, patent families set is used for classification. Since the members of the same patent family disclose the same invention, only a single member of the patent family has been classified.
- **Precision** (or positive predictive value), intended as the fraction of relevant patents among the global patent set. This value should be high enough to grant the proper significance to statistical data retrieved from the set. Precision is evaluated by manually analyzing a sample subset of patents.
- **Level of recall**, which defines the fraction of relevant patents that were included in the set (total number of documents retrieved that are relevant/total number of relevant documents in the database).
- **Innovation Index (I.I.),** a qualitative representation of the information related to the patenting strategies.

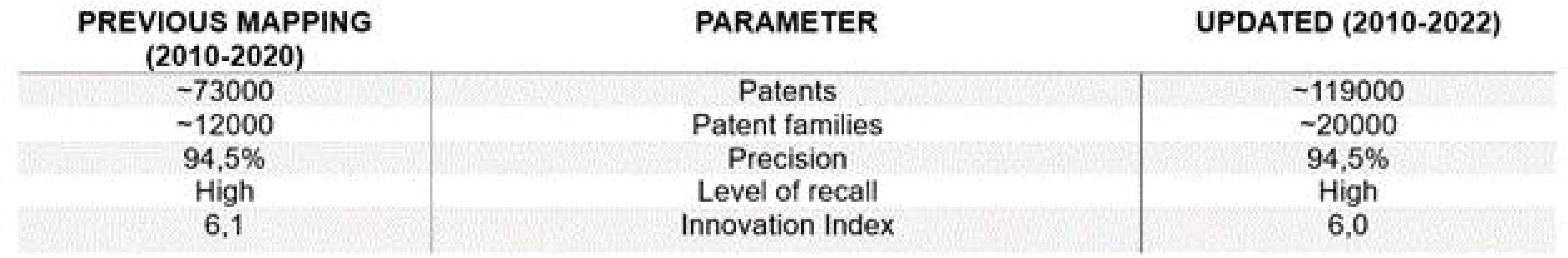

The number is obtained from the ratio between the global set of patents and the bibliographic data (DOCDB [document database]) families of a patent set; the higher the level of extended patents, the higher the value generated by those patents. The number of patent families is indicative of inventive activity and shows how the I.I. category strongly supports the trend to extend patents beyond the area of original invention. Indeed, the number of families can show the spreading of the inventions in different countries.

Despite the slow and steady growth of patent families, there was a significant reduction in the overall number of patents (specific data not provided), leading to a slight decrease in PM I.I. from 6.1 to 6.0.

Figure 2 illustrates the overall trend of the patent families filed (left chart) together with the percentage of the global filed families for filing office (pie plot on the right).

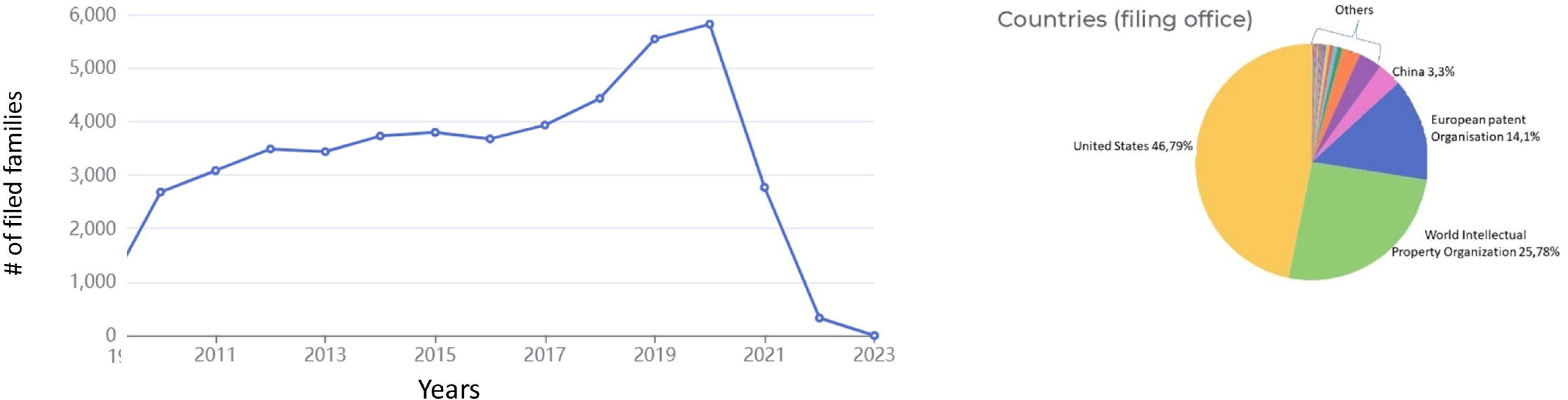

The decrease observed in the chart, related to 2021-2022, is attributed to the 18-month secrecy time of the patents prior to publication.

Having a look to the filing offices, it should be pointed out the presence of the World intellectual property organization (WPO or WIPO)[18] and the European patents organization (EPO)[19]. WPO is a self-funded agency of the United Nations acting as global forum for intellectual property (IP) services, policy, information and cooperation. EPO is an international public organization created by the European Patent Convention that works on national and international patent grants.

Among all the countries, United States retains top spot in global patent activity (46.79%), followed by Europe (14,1%) and China (3,3%). Interestingly, with the respect to previous outcomes, the patenting activity (which can be intended as a primary indicator for innovation in PM) presents a turnaround and it is now lead by universities followed by industries and other categories (i.e., non-profit, hospitals, government, etc.). This also emerges by the assignees analysis wherein the prevalence is pointed out. The top 5 rank shows only the main assignees: the Harvard University (US) followed by University of California (US), Medtronics (Ireland, EU), Standford University (US), University of Texas (US) and Philips (NL) among others.

### EU-China compared statistics

The EU-China compared statistics were obtained by filtering the global analysis; the EU/China attribution is based on the geolocation of the patent family assignees. In the previous work, third countries such as Canada and Israel were included in view of the important links and collaborations already established within the European PM initiatives. These Countries have been maintained in the update analysis.

For this reason, countries that are considered EU are:

- EU members
- European Free Trade Association
- European Economic Area
- Canada
- Israel
- Balkan countries **X** (other than Croatia, Greece)

### Included in STATS

Austria, Belgium, Canada Cyprus, Croatia, Czech Republic, Denmark, Estonia, Finland, France, Germany, Greece, Hungary, Iceland, Ireland, Israel, Italy, Latvia, Lithuania, Liechtenstein Luxembourg, Malta, Netherlands, Norway, Poland, Portugal, Slovakia, Spain, Romania, Slovenia, Sweden, Switzerland, UK

### Excluded from STATS X

Albania, Andorra, Armenia, Azerbaijan, Belarus, Bosnia and Herzegovina, Bulgaria, Georgia, Kosovo, Macedonia, Moldova, Monaco, Montenegro, Russia, San Marino, Serbia, Turkey, Ukraine, Vatican City.

An overview of the statistic is depicted in Figure 3 (table on the top left). Predominance of EU is maintained, but a decrease is experienced since the pick of filed families experienced in 2017 (650 EU, 100 China).

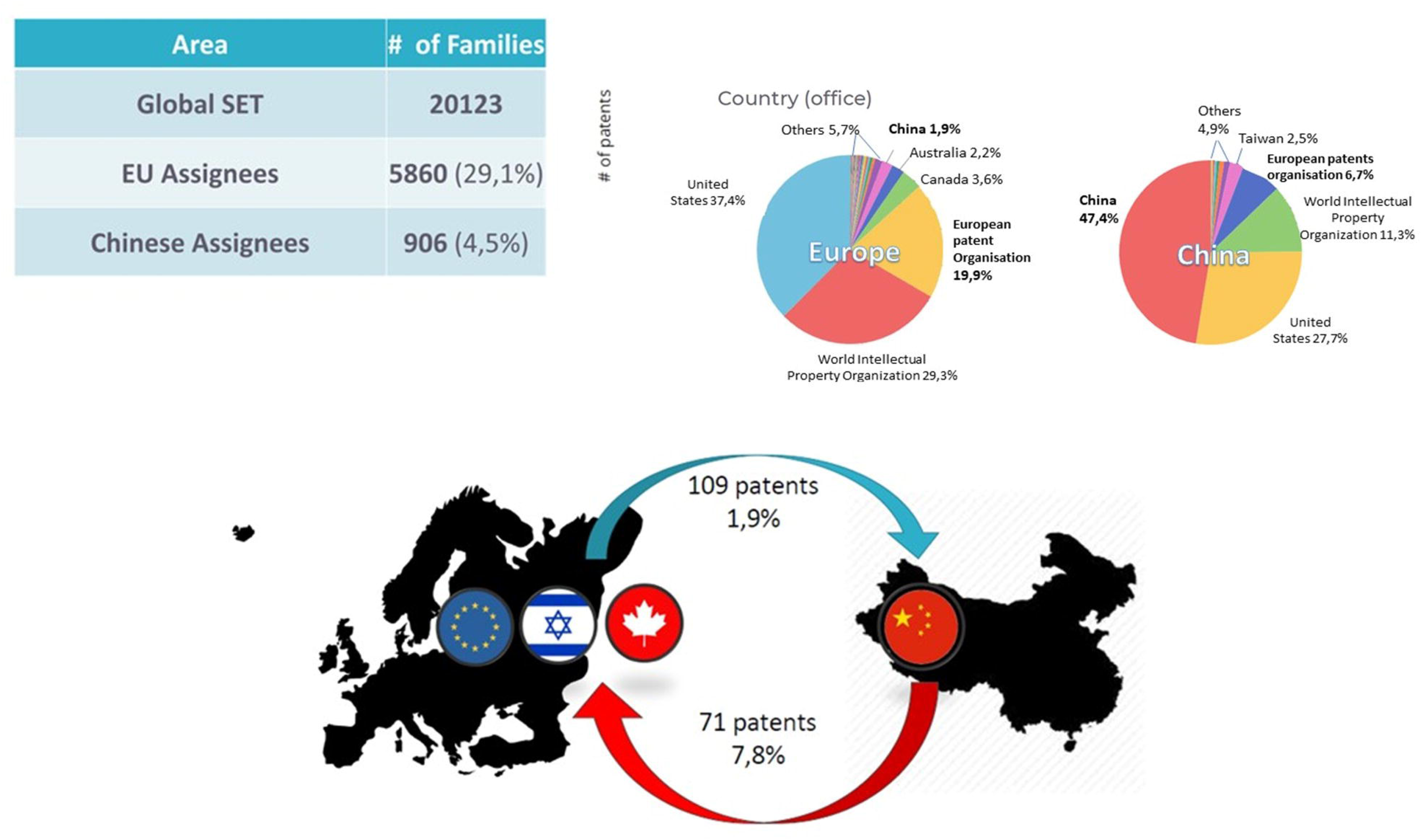

European or Chinese headquarters can file patents by a non-EU/non-Chinese Company or can be coassignee to a non-EU company. Having a look to the main filing offices for Europe and China it emerges that most of the EU patents are filed in US while almost half of the Chinese ones are deposited in China. Below, the percentage of filing of European patent families to China and vice versa. The “filing” analysis is depicted in figure 3). China’s strong interest in opening to the Western market is still evident from the data, while the EU’s entry into the Asian market does not seem to have been stimulated. This delay is still speculated to be due to the difficulty generated by cultural barriers that hinder the relationship with the Chinese market and its rapid expansion [20].

Having a look to the key players in patent filing, companies play a dominant role in both China and Europe, despite the greater presence of academia in China, confirming the results of the previous mapping. However, it is worth noting that many companies in China are state-owned.

Regarding the industry players, the scenario has not changed much. For Europe, the top 5 chart include Philips (NE), Roche (CH), Varian Medical Systems (US) and Siemens (DE) while in China the strong presence of academia is evident, with the University of Hong Kong as the main awardee, followed by Beijing Biothec&Medical Company (Beijing), Enanta Pharmaceutical (US) and the Chinese Academy of Sciences (Beijing).

Collaborations are also analyzed to catch information on the level of collaboration in PM field. It is necessary to distinguish the two definitions as follow:

- ***CO-ASSIGNEED*:** patent having at least 1 Chinese assignee and at least 1 EU assignee
- ***COLLABORATION***: patent of a family having at least 1 Chinese assignee and 1 EU assignee

In this framework, we found a total of **256** collaboration patents from **35** collaboration families, **116 additional documents to the previous dataset**. Data trend is presented in Figure 4. While a strong increase was tracked after 2019, a drop is evident in concomitance to the recent Pandemic that could have impacted collaborations (chart on the left).

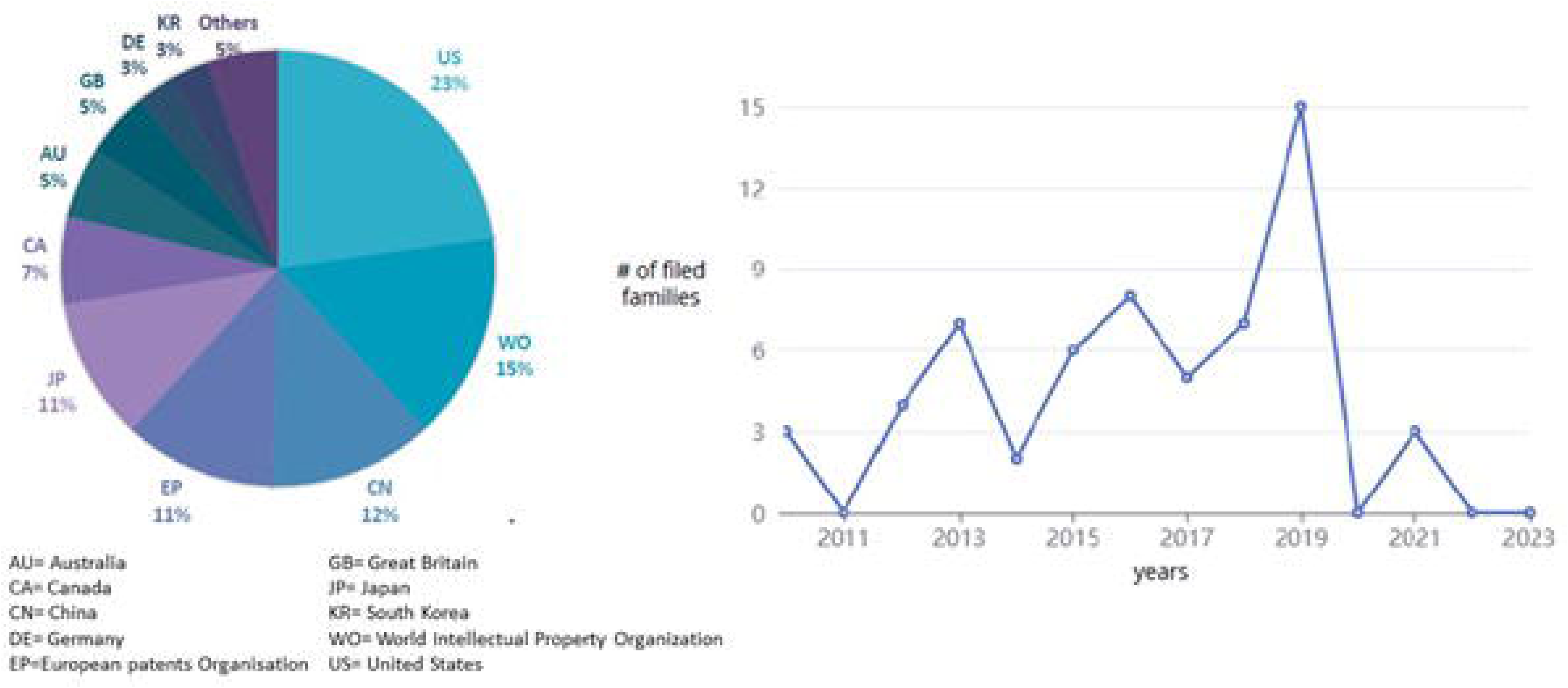

### Patent differential dataset summary 2020-2022

- **Global set of families: 7946**
- **EU Assignees**: 2148
- **Chinese assignees:** 339
- **Collaboration families**: 9
- **Collaboration patents:** 77
- **Patent filing from EU to China**: 3.6%
- **Patent filing from China to EU:** 9.6%

## Mapping methodology: Scientific papers

### Operative Flow

The same strategy used for the previous literature mapping was adopted for the presented update and schematically presented in Figure 5 and summarized below.

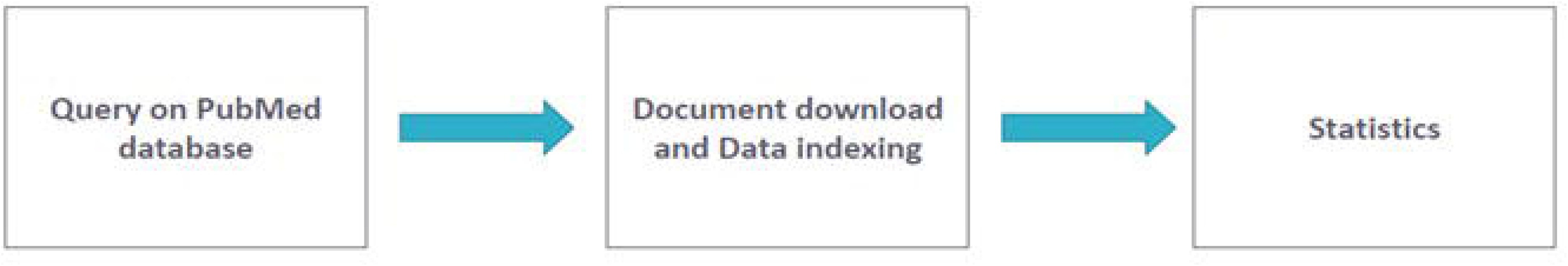

The mined database was PubMed[21], a widely used free online database, provided by the National Center for Biotechnology Information (NCBI), which functions as a container for scientific publications in the medical and life sciences fields. PubMed provides abstracts and links to the full text when available. PubMed exploits MeSH (Medical Subject Headings), the National Library of Medicine (NLM) controlled vocabulary thesaurus used for indexing articles for PubMed, which is a way of improving the accuracy in searching items.

As query strategy, the hybrid approach used for the precedent mapping was carried out merging two different sets

> Set 1) “precision medicine” and “personalized medicine” as MeSH terms

> *Set 2*) *customized query, using keywords*

Set 2 includes non-English written papers only if at least one of title, abstract or keyword is available in PubMed but in English. The filters used were “human” and the time frame from January 2010 to January 2023 (included). January 2023 was included because of some submitted document which indicated a postponed publication date. PubMed database could be partially updated with 2022 and 2023 publications. Additionally, some countries or some journals could provide articles to PubMed with a bigger delay. We obtained a total of **66 950** scientific papers.

A normalization procedure was also necessary. The database may return different names for a single affiliation or express names with different formats. For example, symbols and punctuation were removed and certain words like “center” and “center” were standardized. This procedure allowed for a univocal assignment of a paper to an affiliation, even when the affiliation name is presented in a different format. Nonetheless, the overall number of papers (global set) allocated to each affiliation could be underestimated due to errors in the database or intricate name formats that cannot be controlled.

The results that emerged from the query have been downloaded as .xls format including information about authors, affiliation, year of publication, journal, volume, etc. More precisely, the global paper set has been downloaded in a Medline format and through EndNote (a commercial reference management software used to manage bibliographies and references for scientific publications). The information was then clustered in Global and EU/CN collaborations.

### Global set analysis

The updated mapping results are presented in Table 2.

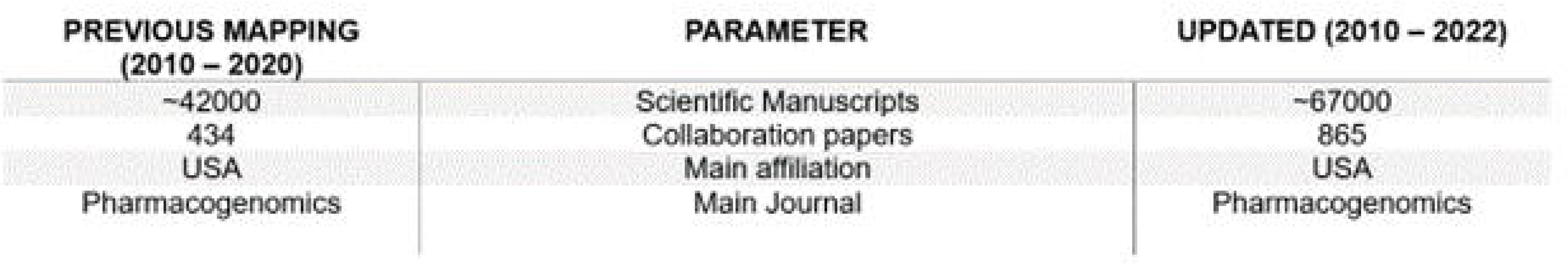

The global analysis data shows a constant growth until a peak in publication in 2019, with around 8000 items. This may depend to the PubMed’s updating practice on which we have little information. Some documents deposited in 2019 were only retroactively uploaded after 2020 and this created an ambiguity that produces the scattered data shown in Figure 6. For this reason, it is necessary to recall the data from latter two years could be underestimated.

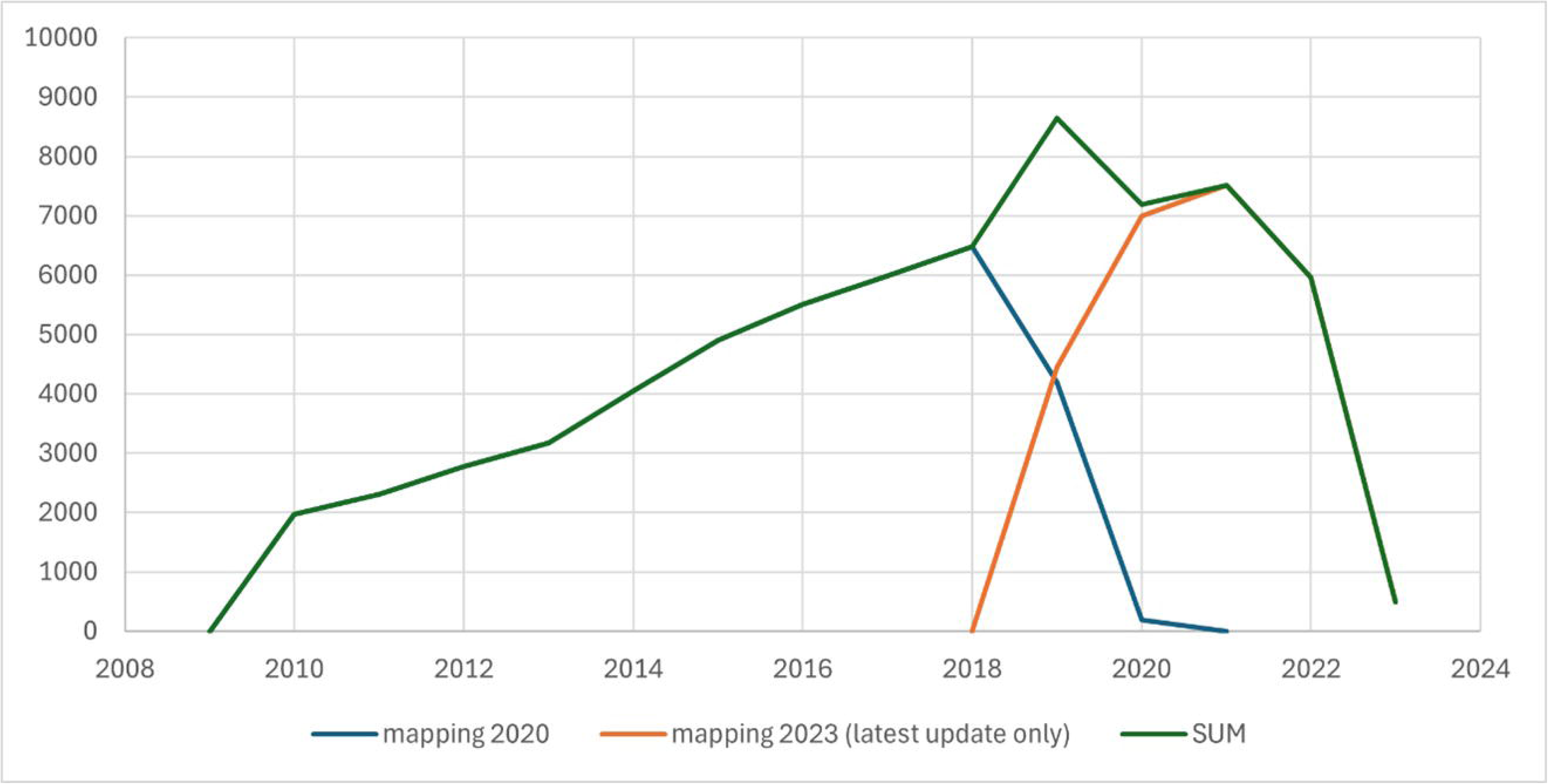

Analysis of assignees was conducted by considering a scientific paper is assigned to a country if at least an affiliation of that country is present. Now the global set presents a total of **66950** publications, **28289** by EU affiliations and **7306** by Chinese. The detailed list of countries is presented in Table 3.

| **RANK** | **COUNTRY** | **N° OF PAPERS** |
| --- | --- | --- |
| **1** | US | 22689 |
| **2** | China | 7306 |
| **3** | United Kingdom | 6120 |
| **4** | Italy | 5007 |
| **5** | Germany | 4627 |
| **6** | Canada | 3384 |
| **7** | France | 3313 |
| **8** | Netherlands | 3264 |
| **9** | Japan | 2579 |
| **10** | Spain | 2575 |
| **11** | Australia | 2381 |
| **12** | Switzerland | 1682 |
| **13** | Sweden | 1439 |
| **14** | Belgium | 1174 |

The information obtained from geolocation, taking into account the PubMed raw data structure, is quite accurate and reliable, while it is more complex to ensure the exact number of publications for the individual legal entity. This bias arises from the different ways in which the same affiliation can be referred to in the DB. The individual name may contain errors and/or omissions, may be used in different combinations (e.g., SinoEUPerMed, SinoEU PerMed, SinoEU-PerMed). For the university, the issue may be more complicated, as the affiliation may contain the university number or include only the department, while others may contain both. In addition, city information may be missing. Fortunately, however, the country is present, so the geolocation of the documents become more reliable to correctly identify the territorial jurisdiction.

On the most active countries in PM literature, the United States maintains its top position, with China securing the second spot, followed by Italy in 3rd place, and the United Kingdom and Germany in the following positions. While the overall landscape remains consistent, slight variations can be appreciated while comparing the rank of top 20 global affiliations (together with the respective total number of deposited manuscripts). The updated information is presented in Table 4, including the respective total number of deposited manuscripts. Currently, 50% of these affiliations are situated in the US, reinforcing its prominence, while China, now positioned 2^nd^, climbed the rankings by doubling its scientific production in the last two years of mapping. Relevant centers are the Department of Clinical Pharmacology from Xiangya Hospital and Central South University in Changsha. The latter has experienced a significant surge in scientific publications on PM, increasing from 151 to 405 papers, consequently advancing three places in the ranking. It is worth noting the absence of normalized data can lead to underestimates in the numbers that are assigned at this stage. Saudi Arabia drops from 1st to 5th place, and both the Netherlands and Japan no longer feature in the ranking. New entries include Germany, Italy, Iran, and Iceland.

| **RANK** | **AFFILIATION** | **COUNTRY** | **# OF PAPERS** |
| --- | --- | --- | --- |
| 1 | Department of Pathology and Genomic Medicine, Houston Methodist Hospital, Houston, Texas. | USA | 452 |
| 2 | Department of Clinical Pharmacology, Xiangya Hospital, Central South University, Changsha, China. | China | 414 |
| 3 | Center for Genomic Medicine, Massachusetts General Hospital, Boston, MA, USA. | USA | 406 |
| 4 | Institute of Clinical Pharmacology, Central South University | China | 405 |
| 5 | Center of Excellence in Genomic Medicine Research, King Abdulaziz University, Jeddah, Saudi Arabia. | Saudi Arabia | 390 |
| 6 | German Cancer Consortium (DKK), German Cancer Research Center (DKFZ), Germany | Germany | 373 |
| 7 | Università Cattolica del Sacro Cuore, Rome | Italy | 355 |
| 8 | Department of Internal Medicine, National Taiwan University Hospital, Taipei | USA | 330 |
| 9 | Department of Biostatistics, Boston University School of Public Health, Boston | USA | 275 |
| 10 | University of Medical Sciences, Tehran, | Iran | 272 |
| 11 | Department of Pathology. The University of Texas MD Anderson Cancer Center, Houston, Texas | USA | 271 |
| 12 | Faculty of Medicine, University of Iceland, Reykjavik | Iceland | 267 |
| 13 | Department of Genomic Medicine, The University of Texas MD Anderson Cancer Center, Houston, Texas | USA | 263 |
| 14 | Department of Molecular and Human Genetics, Baylor College of Medicine, Houston | USA | 257 |
| 15 | Department of Genetics, Stanford University, Stanford, California | USA | 222 |
| 16 | Department of Psychiatry, University of California San Diego, La Jolla, CA | USA | 220 |
| 17 | Department of Psychiatry, University of Toronto, Toronto, ON, Canada. | Canada | 216 |
| 18 | Department of Epidemiology Harvard TH Chan School of Public Health, Boston, MA, USA. | USA | 213 |
| 19 | Department of Psychiatry, Harvard Medical School, Boston, MA, United States | USA | 209 |
| 20 | Wellcome Trust Centre for Human Genetics, University of Oxford, Oxford | UK | 201 |

### EU-China compared statistics

The EU/CN compared statistics were obtained using the same group of EU countries used for the patent mapping (refer to patent section).

The differential global set shows a total of **25418** documents, 45,11% EU affiliations and 15,73% Chinese affiliations. The trend is also depicted in Figure 7 and shows a slight but constant increase in the overall number of published papers. The raw data are filtered to highlight EU and CN affiliations.

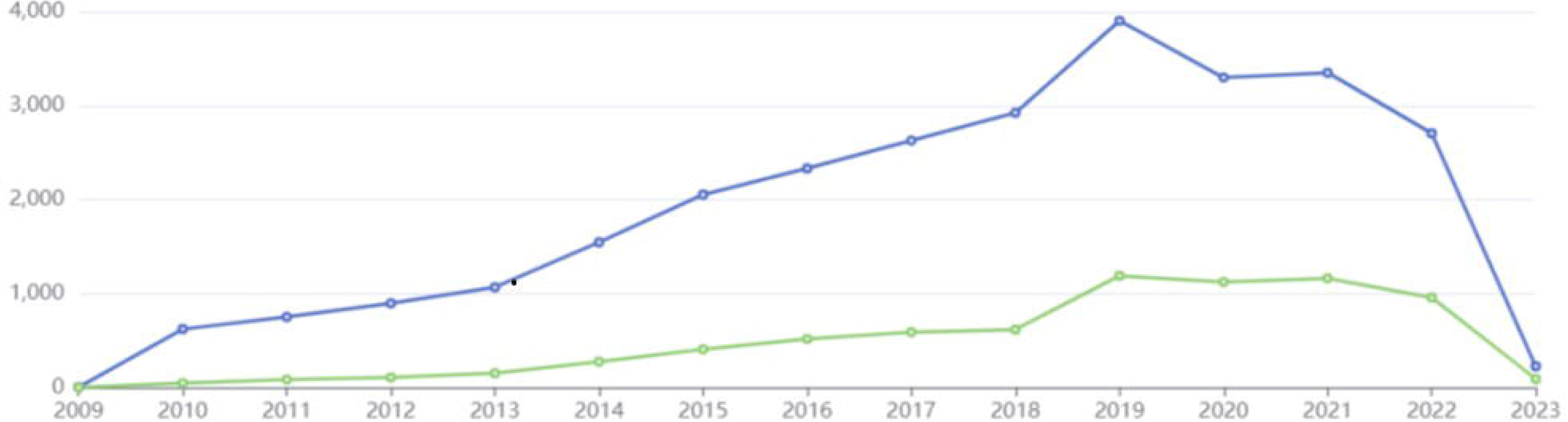

On the top 10 affiliations, several differences to the old mapping can be noted for EU. Germany, Italy and Iceland are in the top three positions overtaking UK from 1^st^ to 5^th^ and Netherlands, now 6^th^ and 8^th^. (Figure 8).

**Figure 8.**
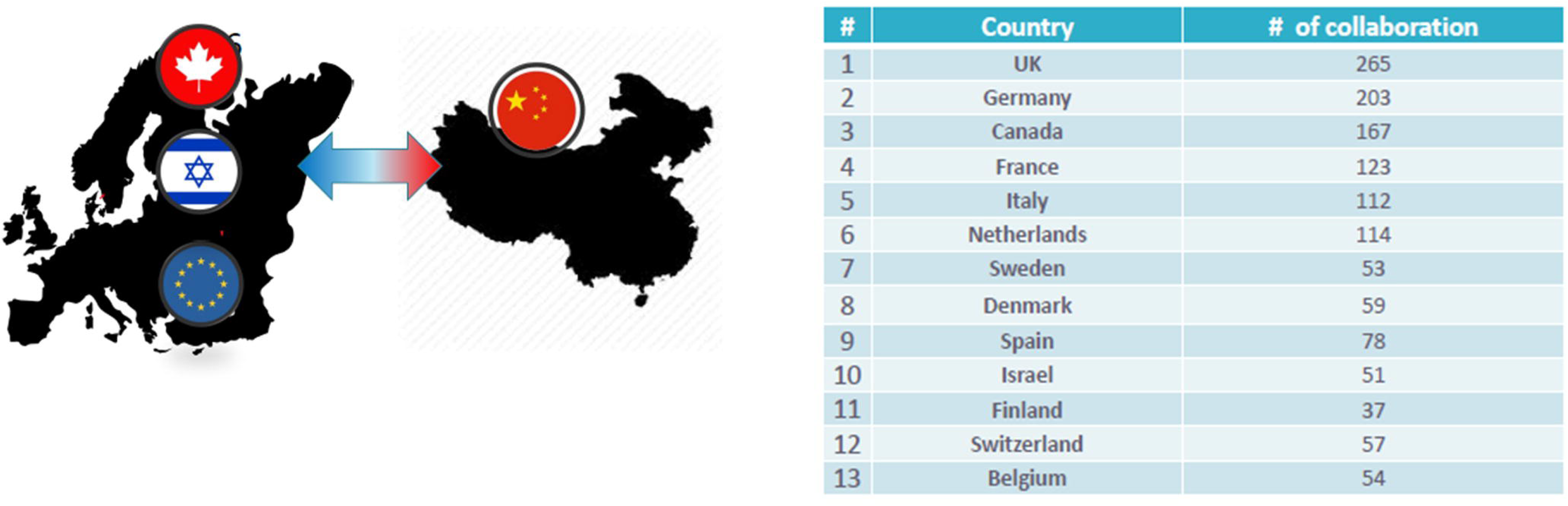
A table containing the main EU Countries who publish in collaboration with China.

In Germany, notable affiliations include the German Cancer Consortium (DKTK) and the German Cancer Research Centre (DKFZ), which contributed 256 new papers. The Università Cattolica del Sacro Cuore, Italy, entered the scene with a total of 355 publications, along with another newcomer, Denmark, which secured the 10th position with the Faculty of Medical and Health Sciences at the University of Copenhagen (164 manuscripts).

Regarding Chinese affiliation, the predominance of the Central South University from Changsha appreciated from the global mapping is well reflected also in this rank. The top 3 is led by Department and Institute of Clinical Pharmacology which, overall, duplicated its publications. The Wenzhou’s Institute of Genomic Medicine (Wenzhou Medical University) drops from 3^rd^ to 4^th^ position, but it presents just 2 new publications. A new entry that is interesting to highlight due to the total high number of publications (84) is The Key Laboratory of Carcinogenesis and Cancer Invasion of the Chinese Ministry.

Collaborations between Europe and China were obtained by considering papers that have at least 1 co-author with a Chinese affiliation and at least 1 co-author with a European affiliation.

A total of **865** collaboration papers (1.3 % of global set) emerged:

- 11.8% of Chinese papers
- 3.0% of European papers

Compared to previous data, even if the percentage of collaboration papers has increased significantly in number (from 434 to 865, with a total of 418 new joint publications during 2020-2022), the scenario is unchanged. EU/CN collaboration papers are the 1,3% of the global set (+0,2%); there is a small drop of Chinese papers percentage (-3,3%) together with a little increase of European papers (0,4%). However, considering the small entity of the variations, any comparison could be speculative. A plot depicting the number of published papers per years, having at least an EU affiliation and at least one Chinese affiliation (Figure 9).

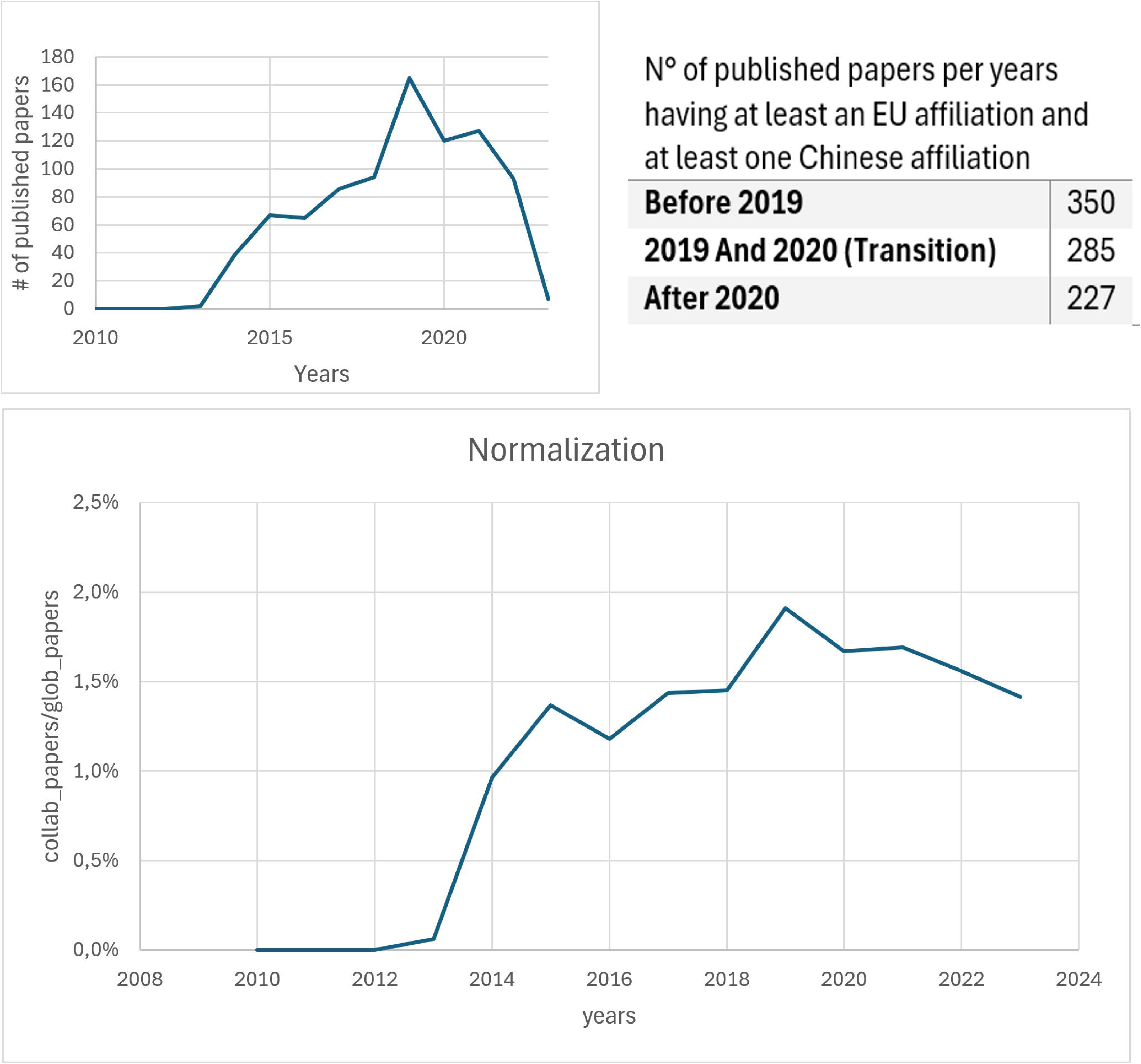

Having a few publications before 2014, the trend constantly grows year by year with a peak in 2019. Going to 2023, there is a smooth decline in the curve trend. This is is due to the underestimation of papers for 2022 derived by postponed publication, and the partial data in 2023. Thus, the number of collaborations continues to be substantial, although at the time this paper is submitted it is not possible to affirm the trend is still growing after 2019. Notably, the ratio with the global number of papers allows to normalize the trend of collaborations against the global trend in PM. From 2015, it remains fairly stable around 1.5%.

About the Scientific journals, many high impact factor Journals are present, such as *Nature* and *PloS one*. The *Journal of Pharmacogenomics* (2436) and *PLoS One* (1035) maintain the first positions while *Scientific reports* has double the number of publications (+405), ranking to the third position with 785 papers. It would be worth noting the list does not present special journals dedicated to PM.

**Scientific papers differential dataset summary 2020-2022**

- **Global set of scientific publication:** 25418
- **EU Assignees**: 11467
- **• Chinese assignees:** 3999
- **Collaboration papers**: 418 (1.6% of global set, 10.4% of EU set, 3.6% of CN set)

### Mapping of pre-prints

Peer-review is a crucial process, essential for assessing the integrity and consistency of scientific papers, a mandatory prerequisite for publication in scientific journals, regardless of their prestige. Although it is unavoidable, the peer-review process can often lead to significant delays in publications due to various biases and problems, although discussion of these challenges is beyond the scope of this work. However, this hampers scientific communication, prompting researchers to seek alternative strategies for faster dissemination of their results. This need has fueled the adoption and proliferation of non-peer-reviewed papers, also known as preprints, accepted as a valid dissemination practice. Preprints have become familiar in various fields of research since the early 1990s, particularly in physics, with the creation of the first arXiv.org server in 1991. Since the last decade, life sciences researchers also had back of special dedicated databases, which growth was boosted during the recent pandemic because of the urgency of sharing potentially benefitting results as quick as possible [22].

In line with the workflow strategy outlined for papers, the mining of two dedicated databases, namely bioXriv[23] and medXriv [24], was included in the update. The objective was to retrieve the possible slice of literature of interest that has not yet been published but is present in the form of preprints. It must be borne in mind that the lack of experts/referees’ oversight on preprints can affect their integrity, necessitating critical use.

A total of **1682** items on PM were collected. Unfortunately, due to the difficulties of handling this raw data, it was not possible to perform accurate statistics. Nevertheless, the results are included in the web platform and accessible for consultation.

### SINO-EU PerMed web platform

The data obtained from the mapping update and shown in this work is available on the project online platform. Through this tool, which is interactive, it is be possible to access and consult the complete information or filter data according to chronological criteria, geolocation or medical topic addressed. The database is accessible through a dedicated section of the project web page[10] and it is achievable from the link https://www.sino-eu-permed.eu/en/Database-1775.html [25].

The database offers customizable data visualizations, including all the graphs presented in this document, and can be downloaded in .xls and .jpg formats. Indeed, most of the plots presented in this manuscript were obtained by the database. For both paper publications and patent documents the web platform is provided with search filter (goal, medical field, tools, stratification approach, country) and other query tools, in order to help the researchers to select quickly and accurately the documents subset to be analyzed. Further, the embedded module could be an effective solution for extracting customized statistical data. The project encourages the community to use this resource enhancing their understanding of the Sino-EU collaborative landscape in PM and contributing to the collective knowledge and growth of the international PM community.

## Conclusions

The updated mapping offers a comprehensive overview of Sino-European collaborations in the field of PM. Spanning from 2010 to 2022, the data reveals a sustained level of collaboration, evident in both patent filings and the production of scientific publications. The involved stakeholders remain consistent, with companies playing a predominant role in patent filing for both China and Europe, although academic involvement is higher in China, aligning with the prior mapping findings.

A noteworthy observation from the latest update is the substantial increase in collaborations, as reflected in dedicated paragraph of this paper. By examining the differential data, there is a notable rise in collaborative patent filings (77 items) compared to the previous decade (140 identified during 2010-2020). Similarly, the number of collaborative papers in the last biennium matches the quantity mapped in the 2010-2020 decade (418 new items compared to 434 previously emerged).

This surge in collaborations could be attributed to the needs arising from the historical period, in particular to address the challenges posed by SARS-CoV-2 and the urgency to mitigate its consequences. Although the inclusion of the 2020-2022 period provides a preliminary view of the impact of the Covid-19 pandemic on Sino-EU and global PM collaborations, it is crucial to recognize the blind periods associated with patents and the time between peer-review and article publication processes, factors that influence the temporal analysis presented. Despite efforts to mitigate bias by including preprints in the mapping process, this did not contribute significantly to the analysis of pandemic effects. However, the appearance of 1682 preprints underline the substantial interest and engagement surrounding the PM topic within academia.

It is imperative to consider the speculative nature of these claims, acknowledging the need for further statistical investigations to support insights derived from the mapping data. While the data obtained serves as a relevant pool for future exploration and more sophisticated statistical analyses, a comprehensive understanding of collaborative dynamics necessitates not only an investigation of the existing partnerships but also a thorough examination of the ethical and legal dimensions that may potentially hinder such synergies. Future efforts will focus on implementing an extended approach that integrates studies focusing on ethical considerations and legal frameworks, essential for fostering sustainable partnerships in this evolving field.

## Data Availability

data presented in this manuscript are freely accessible online through the dedicated database

https://tls.consult.errequadrosrl.com/

